# Building accessible resources to empower communities: the case of the Lupus Mexican Registry

**DOI:** 10.64898/2026.06.11.26355300

**Authors:** Domingo Martínez, Driselda Sánchez-Aguirre, Grecia Sevilla-Parra, Itzel Olivares-Martínez, Fernanda Bravo-García, Ana Laura Hernández-Ledesma, Luis A. Aguilar, César Arturo Dominguez-Frausto, Jair García, Angélica Peña-Ayala, Deshiré Alpízar-Rodríguez, Lizbet Tinajero-Nieto, Sarael Alcauter, Alejandra Medina-Rivera

## Abstract

**Motivation:** Although SLE data in Latin America is increasing, clinical datasets remain difficult to access and interpret, highlighting the need for accessible tools that support data-driven precision medicine, citizen science, and public health initiatives.

**Results:** We developed a user-friendly platform that enables us to explore LupusRGMX data through interactive queries, report generation, statistical modeling, and comprehensive insights. This resource supports community-oriented research, improves the visibility of underrepresented populations in lupus research, and provides a useful tool to enhance data accessibility.

**Availability and implementation:** Developed in R using Shiny and bslib for interactive visualization and interface design. Available at https://github.com/NeuroGenomicsMX/Lupus_App_2.0 and https://lupusrgmx.liigh.unam.mx/shiny/lupus/

## 1. Introduction

Systemic lupus erythematosus (SLE) is a multifactorial autoimmune disease with a wide spectrum of clinical manifestations affecting multiple organs [1]. Epidemiological studies have consistently reported differences in disease prevalence, incidence, severity, and mortality across populations, reflecting the complex interplay between genetic and environmental factors contributing to this variability [1,2]. In addition, a significant gap persists in the representation of populations from low and middle-income countries, limiting the generalizability of the current knowledge and contributing to disparities in disease understanding, diagnosis, and management [1,2].

Latin America comprises diverse populations shaped by distinct socioeconomic, genetic, environmental, and historical contexts, which are reflected in the variability of SLE prevalence, estimated to range from 34.9 to 91.9 cases per 100,000 individuals [3]. Several large-scale efforts, including the Grupo Latinoamericano de Estudio de Lupus, the Almenara Lupus cohort, the LUMINA cohort, and the Mexican Lupus Registry, have contributed to characterizing disease patterns across the region [4-7]. These studies highlight the role of genetic ancestry and environmental exposures in shaping disease heterogeneity. However, persistent challenges, including limited epidemiological data, high healthcare costs, shortages of rheumatologists, and insufficient disease awareness, continue to drive disparities in disease outcomes and access to healthcare [3].

Consequently, despite substantial progress in characterizing SLE across the region, a comprehensive understanding of SLE in Latin American populations remains incomplete. Moreover, while clinical registries are increasingly available, their accessibility and usability are often restricted, particularly for clinicians and researchers without computational expertise, as well as patients, associations, caregivers and family members, and public-health policymakers. This gap underscores the need for open, interactive, and user-friendly tools that empower communities to efficiently explore complex clinical data and facilitate knowledge translation.

Here, we present an accessible interactive platform based on the Mexican Lupus Registry (LupusRGMX), designed to facilitate the exploration, modeling, and interpretation of complex clinical and epidemiological data. Built using Shiny [8], this tool enables clinicians, researchers, and other stakeholders to dynamically query and visualize population-level insights without requiring advanced computational skills. By improving data accessibility and interpretability, the platform supports hypothesis generation, promotes data-driven decision-making, and enhances the visibility of underrepresented populations. Importantly, such tools may also contribute to broader community engagement and inform public health strategies aimed at reducing disparities in disease management and access to care.

## 2. Methods

### 2.1. Data source

Data were obtained from the LupusRGMX cohort [7,9], an electronic registry dedicated to Mexican patients diagnosed with Systemic Lupus Erythematosus (SLE). This research protocol was approved by the Ethics Research Committee of the Neurobiology Institute at the National Autonomous University of Mexico (approval no. 093.H). The study population included patients aged 18 years or older with a confirmed diagnosis of SLE. Initial data curation steps involved the removal of duplicate records and the exclusion of variables exhibiting missing values, reaching a final dataset of 895 cases with 73 clinical and epidemiological variables.

### 2.2. App architecture

The interactive web application was developed in R [10] using the Shiny framework [8] to provide dynamic user interactions. The application’s interface design and theming were constructed using the bslib package [11]. To optimize performance and maintainability, the architecture relies on a modular design divided into distinct logical tabs, employing a progressive disclosure strategy to prevent interface overload and present detailed results only when requested. Data manipulation and structuring within the application are handled primarily through the tidyverse ecosystem [12].

### 2.3. Data processing

As a first stage, a user-needs assessment survey was conducted via Google Forms to identify the core functionalities required for the application. The survey gathered 35 responses from a diverse group of stakeholders, including leaders of patient associations, rheumatology specialists, patients, and the public.

Next, for the clinical relevance and analytical utility of the application, a committee of six experts conducted a variable selection process using a consensus-based approach, resulting in a final set of 73 variables. Such variables were categorized into distinct domains: clinical variables, sociodemographic factors, patient-reported outcomes (PROMs), lifestyle and quality of life, and neurolupus markers. To ensure traceability and reproducibility across all project stages, original REDCap variable names, including checkbox encoding formats, and participant identifiers (record_id) were strictly preserved. It is worth mentioning that findings from this user-needs assessment were shared with the committee of six experts to guide their discussion process.

Missing data patterns were evaluated using visualization techniques without excluding subjects. For the final application dataset, a completeness threshold was established, retaining only variables with fewer than 1500 missing values, resulting in a total of 895 cases. Within the application’s environment, all variable handling and data curation occur locally in the script immediately preceding the execution of each analytical module. While internal variable names remain immutable, outputs are translated into human-readable Spanish concepts to enhance user interpretability within the target audience. A secondary processing stage integrates the neurolupus sub-dataset, comprising MRI findings, VolBrain metrics, and MoCA assessments.

### 2.4. Features

The platform is functionally organized into five primary modules: General Report, Comparisons, General Statistical Modeling, NeuroLupus Statistical Modeling, and Raw Data Access (Fig. 1). Because of the target user, all texts are shown in Spanish on the app. However, from now on, all figures are shown in English via the automated translation made by the Google Translate Web tool.

**Figure 1.**
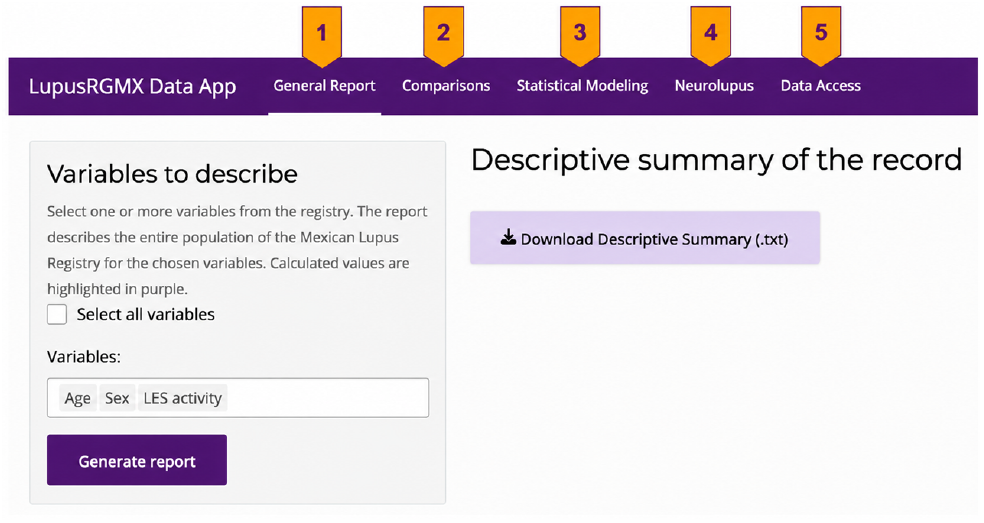
Main user interface and application architecture. The platform is organized into five core modules (labeled 1–5): General Report, Comparisons, General Statistical Modeling, NeuroLupus Statistical Modeling, and Raw Data Access.

#### General Report

Generates dynamic descriptive statistical summaries based on user-defined general filters, such as age, sex, and comorbidities.

#### Comparisons

Facilitates dynamic cross-variable analysis to identify statistically significant associations between patient subgroups and clinical outcomes. The module incorporates an automated routing system that selects the appropriate mathematical approach based on data type, then models or tests assumptions.

General Statistical Modeling: Provides an interface for configuring and executing statistical models, including Linear Regression, Binomial Regression, and Multinomial Regression. Users can select predictor variables to evaluate target outcomes such as SLEDAI, quality of life, or lupus nephritis.

#### NeuroLupus Statistical Modeling

A specialized module that provides another interface for configuring and running statistical models for neurocognitive and imaging outputs by analyzing associations between clinical scores and magnetic resonance findings, such as tract lesions, cerebrovascular events, and MoCA scores.

#### Raw Data Access

A dedicated portal that addresses patient privacy and provides a formal REDCap request form for communities and researchers seeking access to the raw database for collaborative analysis or highly detailed reports.

To maximize accessibility for the target demographic, all interface elements and generated explanations are presented in Spanish, prioritizing clear, non-technical text over complex or technial descriptions.

## 3. Results

The platform processes complex data to return textual reports in human language, prioritizing clinical and conceptual interpretability over statistical technicalities. This allows patient communities, researchers, and healthcare personnel to quickly identify epidemiological insights within the Mexican population without the need for external analysis software. Optionally, complete technical reports or supporting graphs can be downloaded in txt, or PNG format, respectively, by an action button.

### 3.1 General Report

The General Report module enables the generation of dynamic descriptive statistical summaries based on the final dataset of 895 selected cases. Through an interactive interface, users can filter and explore among the 73 clinical and epidemiological variables. To demonstrate the module’s exploratory capabilities, key indicators of familial aggregation and accumulated organ damage (kidney) were analyzed (Fig. 2).

**Figure 2.**
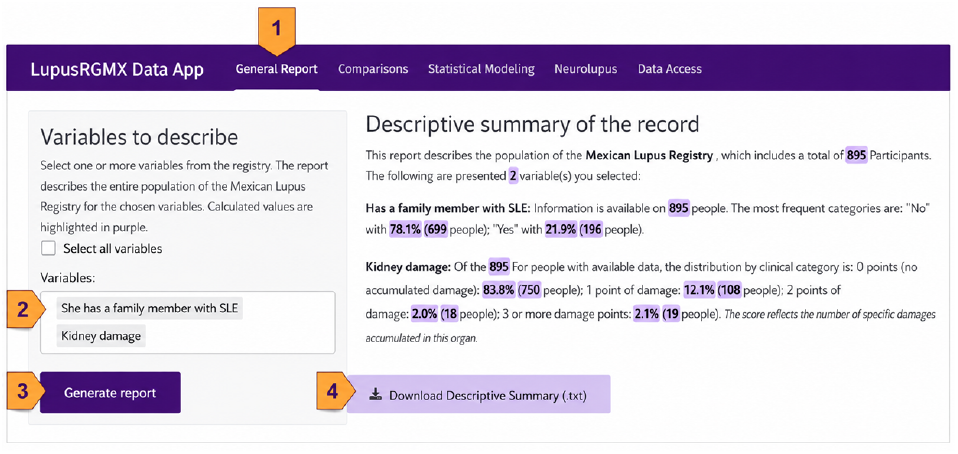
Workflow of the General Report module. The numbered steps (1–4) outline the report generation process: 1) navigating to the module via the main menu, 2) selecting specific variables from the dynamic input area, 3) executing the “Generate Report” action to view the descriptive summary on the right panel, and 4) downloading the report in txt format.

To replicate this illustrative case, users would follow four steps: (1) select the module from the top navigation bar, (2) pick the desired variables from the dynamic selection menu, in this case, has-a-family-member-with-SLE and kidney damage, and then (3) click the “Generate Report” button to render the output in the main viewing area. Finally (4) downloading the txt report. The output looks like this.

#### Familial Aggregation

Among patients with complete records regarding family history (n = 895), 21.9% (n = 196) reported having at least one family member diagnosed with SLE, while the remaining 78.1% (n = 699) reported no family history of the disease.

#### Renal Damage

Among individuals with available data (n = 895), the majority of patients presented no accumulated kidney damage, with 83.8% (n = 750) scoring 0 points. In contrast, 12.1% (n = 108) presented 1 point of accumulated damage, 2.0% (n = 18) presented 2 points, and 2.1% (n = 19) presented 3 or more points of kidney damage. The score reflects the number of specific damages accumulated in this organ.

### 3.2 Comparisons

The comparatives module incorporates an automated routing system that selects the appropriate mathematical approach based on data type. Nominal comparisons are evaluated using Pearson’s Chi-squared tests of independence—reinforced by strict assumption validation to abort calculations if expected cell frequencies fall below 5—and visualized through grouped bar charts detailing absolute frequencies. For continuous or discrete ordinal variables, the algorithm executes robust asymptotic permutation tests via Monte Carlo simulations, followed by post-hoc pairwise comparisons with False Discovery Rate (FDR) correction using the coin package [13]. The interface conditionally renders 100% stacked proportional bar charts for ordinal scales (such as discrete organ damage scores) and smooth density plots for continuous distributions using viridis color scales [14], allowing users to explore clinical interpretation reports and high-resolution (300 dpi) graphical outputs.

To illustrate how to compare continuous variables among groups, age and eating-habits were utilized (Fig. 3).

**Figure 3.**
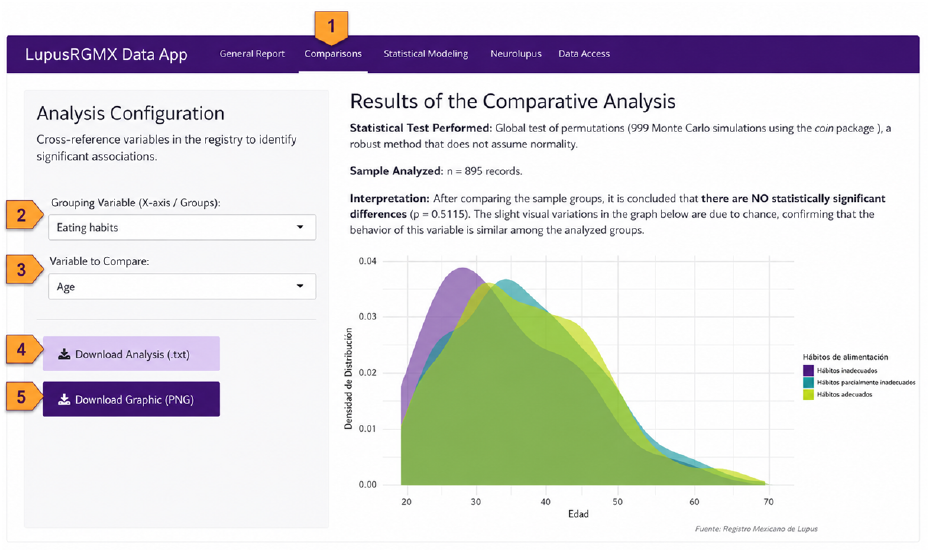
To execute a comparative analysis, users follow a straightforward five-step workflow: (1) navigating to the “Comparisons” module via the top navigation bar, (2) selecting the categorical grouping variable to define the groups, (3) choosing the continuous specific variable to compare across those groups, (4) downloading the automated statistical report, and (5) clicking the “Download Graphic” button to export the high-resolution figure.

To replicate such analysis, users would follow five steps: (1) select the Comparisons module from the top navigation bar, (2) choose the grouping variable, (3) pick a variable to compare, (4) download the report, and (5) click the “Download Graphic” button to get the figure.

Without assuming a normal data distribution, a robust global permutation test using 999 Monte Carlo simulations was conducted using the coin package [13] across the eating habit profiles (n = 895). The analysis revealed no statistically significant differences in age among the distinct groups (p = 0.5115).

To illustrate the discrete variable comparisons among groups, corticosteroids use and SLE activity were used (Fig. 4).

**Figure 4.**
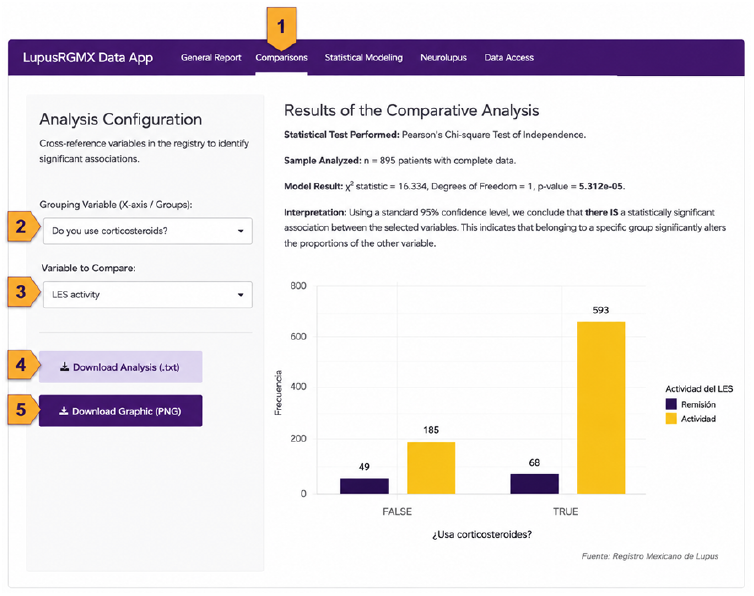
Workflow of the Comparisons module. The platform guides users through a five-step analytical process, annotated in the image: (1) selecting the module from the top navigation bar; (2) choosing the grouping (e.g., corticosteroid use); (3) selecting the variable to compare (e.g., SLE activity); (4) downloading the automated statistical report in plain text format; and (5) exporting the high-resolution graphic. The right panel displays the corresponding dynamic results, illustrating in this case a Pearson’s Chi-square test and a grouped bar chart with absolute frequencies.

To replicate the illustrative analysis, users would follow the steps labeled 1 through 5. (1) Accessing the main module, (2) selecting the grouping variables, in this case, corticosteroids use, and then (3) picking the variable to compare among groups, in this example, SLE activity. Finally, click (4) and (5) to download the text report and graph, respectively.

The on-screen example demonstrates the evaluation of the association between corticosteroid use and SLE activity, displaying the automated statistical output and its visual representation. This analysis demonstrated that, using a standard 95% confidence level, there is a statistically significant association between SLE activity and corticosteroid use (n=895, χ^2^ statistic = 16.334, Degrees of Freedom = 1, p-value = 5.312e-05).

### 3.3 General Statistical Modeling

The General Statistical Modeling module provides a versatile interface for configuring and executing statistical analyses, seamlessly adapting to Linear, Binomial, and Multinomial regressions based on the nature of the selected target variable. Users can evaluate complex outcomes such as disease activity (SLEDAI), quality of life, or organ-specific damage using up to 10 simultaneous predictors.

To demonstrate the platform’s analytical robustness and reproducibility, we used the module to evaluate the determinants of quality of life in the final dataset of 895 selected cases (Fig. 5).

**Figure 5.**
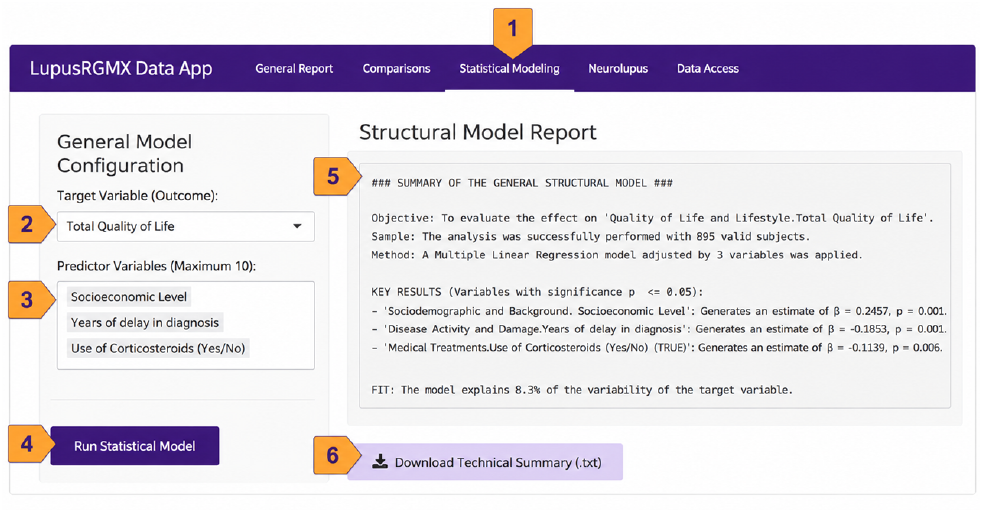
Workflow of the General Statistical Modeling module. The numbered steps (1–6) outline the process for generating a statistical report: (1) navigating to the module via the main menu; (2) selecting the target variable; (3) choosing the predictor variables; (4) executing the model; (5) viewing the comprehensive model results; and (6) downloading a descriptive summary.

A multiple linear regression model was configured with ‘Total Quality of Life’ as the target variable. To replicate this illustrative case, users would follow the next steps. Navigating to the module (1), defining the target outcome (2), in this case, Total Quality of Life; then selecting the predictor variables (3), in this case, socioeconomic status, delayed diagnosis, and use of corticosteroids, and executing the analysis (4). The comprehensive results are then displayed on the right panel (5). Optionally, advanced users can download a detailed technical report for further analysis (6).

The automated text-generation engine successfully identified socioeconomic status (β = 0.09, p < 0.001), delayed diagnosis in years (β = -0.67, p < 0.001), and the use of corticosteroids (β = -2.00, p = 0.01) as significant predictors, with the overall model explaining 8.0% of the variance. These dynamically generated results perfectly replicate recent peer-reviewed epidemiological findings in the Mexican SLE population (9), highlighting the application’s capacity to yield accurate, publication-ready statistical insights without requiring external computational software.

### 3.4 Neurolupus Statistical Modeling

The Neurolupus Statistical Modeling module was specifically designed to bridge the gap between clinical assessments and advanced neuroimaging findings, allowing researchers to explore complex associations between cognitive performance, structural brain changes, and systemic disease markers. To rigorously accommodate the underlying topology of the clinical data, the module dynamically selects the appropriate mathematical framework based on the nature of the dependent variable. Multiple Linear Regression is applied to continuous outcomes (e.g., neurocognitive scores or volumetric measurements) to estimate the linear coefficients and effect sizes of multiple systemic predictors. Conversely, Binomial Logistic Regression is utilized for binary endpoints (e.g., the presence or absence of specific morphological anomalies) to provide probabilistic risk estimations via Odds Ratios. This dual-model architecture ensures that both magnitude and likelihood are accurately captured across diverse neuroclinical profiles. To demonstrate this functionality, we analyzed the structural MRI outcome “Total LES Lesion Count” in a specialized sub-cohort of 41 scanned patients (Fig. 6).

**Figure 6.**
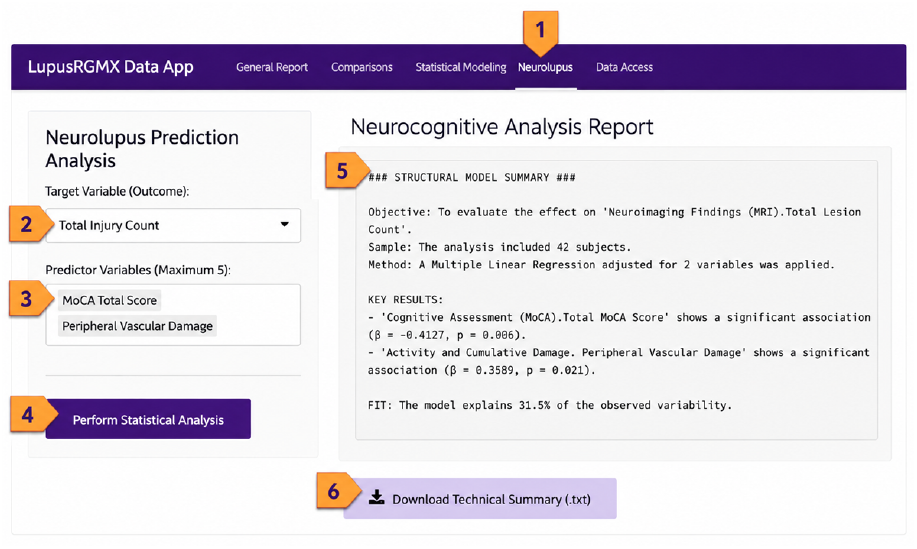
Workflow of the NeuroLupus Statistical Modeling module. The numbered steps (1–6) outline the process for generating a statistical report: (1) navigating to the NeuroLupus module via the main menu; (2) selecting the neurolupus target variable; (3) choosing predictor variables; (4) executing the NeuroLupus statistical model; (5) viewing the comprehensive results; and (6) downloading a wide descriptive summary.

To replicate this illustrative case, users can follow the next steps. Navigating to the module via the main menu (1); selecting the number of brain lesions as the target variable (2); picking MoCA score and peripheral vascular damage as the predictor variables (3); executing the statistical model (4); viewing the comprehensive results on the main panel (5); and (6) downloading a detailed descriptive summary.

Thus, using a multiple linear regression model, the automated system identified a significant inverse association with cognitive performance, where a higher Total MoCA score predicted a lower lesion burden (β = -1.21, p < 0.01). Furthermore, systemic vascular involvement was shown to be a critical factor; the presence of peripheral vascular damage was significantly associated with a higher number of brain lesions (β = 7.04, p = 0.01). Overall, the configured model explained 31.5% of the variance in the total lesion count. These dynamically generated insights underscore the platform’s capacity to facilitate advanced, multidisciplinary research in neuropsychiatric SLE, without the steep learning curve typically associated with specialized statistical software.

## 4. Discussion

Although the platform demonstrated robust performance for exploratory analysis and automated statistical reporting, several limitations should be acknowledged. First, the registry is based on a Mexican SLE cohort, which may limit the generalizability of findings to other populations with different genetic, environmental, and healthcare contexts. Second, despite the implementation of data curation strategies, some variables contained substantial missingness, particularly in specialized modules such as NeuroLupus, where sample size remained limited.

In addition, the automated statistical outputs are intended to facilitate hypothesis generation and data exploration rather than replace formal expert-driven statistical interpretation. Nevertheless, the platform successfully reproduced previously published epidemiological findings and enabled the integration of multidimensional clinical, sociodemographic, neuroimaging, and cognitive data into an accessible environment for clinicians, researchers, and patient communities. These findings highlight the potential of user-friendly, community-oriented computational tools to reduce technical barriers in lupus research. By democratizing access to complex epidemiological data, the platform fosters a citizen science framework that empowers patients and the general public to transition from passive subjects to active stakeholders. Ultimately, this approach improves the visibility of underrepresented populations and promotes collaborative, data-driven strategies in precision medicine and public health initiatives.

## Acknowledgements

This work received support from Alejandro León of the Laboratorio Nacional de Visualización Científica Avanzada. We also thank Carina Uribe Díaz, Christian Molina-Aguilar, and Alejandra Castillo Carbajal for their technical support. Authors would like to express their special acknowledgment to Fundación Proayuda Lupus Morelos A.C, Lupus MX, El despertar de la Mariposa, Realidad Lúpica, Red de Apoyo Somos Valientes Toluca, Abraza con amor, A.C. Enfermedades Reumáticas y Autoinmunes, Aprendiendo a Vivir con Lupus y Fibromialgia A.C., Lupus y Aij Monterrey, and the Centro de Estudios Transdisciplinarios Athié-Calleja por los Derechos de las Personas con Lupus A. C., for their invaluable support. We also acknowledge the support of the MexOMICS Consortium research group and the clinicians who participated in data collection and validation.

## Data availability

The raw data underlying this article are safeguarded to protect participant privacy. Communities and researchers wishing to access the database for collaborative analysis must complete a formal request form via REDCap, which is accessible directly through the ‘Raw Data Access’ module within the application. The source code and ongoing development of the application are publicly available on GitHub at https://github.com/NeuroGenomicsMX/Lupus_App_2.0

